# Organizing Pneumonia of COVID-19: Time-dependent Evolution and Outcome in CT Findings

**DOI:** 10.1101/2020.05.22.20109934

**Authors:** Yan Wang, Chao Jin, Carol C. Wu, Huifang Zhao, Ting Liang, Zhe liu, Zhijie Jian, Runqing Li, Zekun Wang, Fen Li, Jie Zhou, Shubo Cai, Yang Liu, Hao Li, Zhongyi Li, Yukun Liang, Heping Zhou, Xibin Wang, Zhuanqin Ren, Jian Yang

## Abstract

**Objective:** As a pandemic, a most-common pattern resembled organizing pneumonia (OP) has been identified by CT findings in novel coronavirus disease (COVID-19). We aimed to delineate the evolution of CT findings and outcome in OP of COVID-19.

**Materials and Methods:** 106 COVID-19 patients with OP based on CT findings were retrospectively included and categorized into non-severe (mild/common) and severe (severe/critical) groups. CT features including lobar distribution, presence of ground glass opacities (GGO), consolidation, linear opacities and total severity CT score were evaluated at three time intervals from symptom-onset to CT scan (day 0-7, day 8-14, day>14). Discharge or adverse outcome (admission to ICU or death), and pulmonary sequelae (complete absorption or lesion residuals) on CT after discharge were analyzed based on the CT features at different time interval.

**Results:** 79(74.5%) patients were non-severe and 103(97.2%) were discharged at median day 25 (range, day 8-50) after symptom-onset. Of 67 patients with revisit CT at 2-4 weeks after discharge, 20(29.9%) had complete absorption of lesions at median day 38 (range, day 30-53) after symptom-onset. Significant differences between complete absorption and residuals groups were found in percentages of consolidation (1.5% vs. 13.8%, *P*=0.010), number of involved lobe >3 (40.0% vs. 72.5%, *P*=0.030), CT score >4 (20.0% vs. 65.0%, *P*=0.010) at day 8-14.

**Conclusions:** Most OP cases had good prognosis. Approximately one-third of cases had complete absorption of lesions during 1-2 months after symptom-onset while those with increased frequency of consolidation, number of involved lobe>3, and CT score >4 at week 2 after symptom-onset may indicate lesion residuals on CT.

## Introduction

Since late December 2019, the ongoing outbreak of Coronavirus Disease 2019 (COVID-19) related pneumonia, caused by a novel coronavirus, severe acute respiratory syndrome coronavirus 2 (SARS-CoV-2; previously known as 2019-nCoV), has rapidly expanded throughout worldwide (1-3). By 29 April 2020, a total of 3.01 million patients with confirmed COVID-19 pneumonia and 207,973 deaths have been reported (4). Clinical and radiological characteristics of COVID-19 pneumonia have been systematically described. It is noting that the most common findings on chest computed tomography (CT), i.e. peripheral ground glass opacity (GGO), consolidation or both predominantly in bilateral and multifocal distributions highly resembled to a CT pattern of organizing pneumonia (OP) (5,6). As a common lung injury, most cases of OP were demonstrated to have a good prognosis, while permanent damage and interstitial fibrosis were still observed in scare severe cases (7). Similar prognosis was observed in COVID-19, i.e. above 80% of cases had been discharged with recovery (8). Despite this, prognosis of OP pattern in COVID-19 including radiological outcome and disease course relating to resolution of pulmonary lesions remains currently unclear.

A plenty of studies have explored the evolution of pulmonary lesions based on chest CT (9,10). As the disease progresses, increased number, extent and density of GGOs on CT have been observed (11). Among these, consolidation was considered as an indication of poor prognosis (12). However, evolutions of OP pattern in COVID-19 and the relations with radiological outcome have not been well described. This study therefore aimed to delineate the time-dependent evolution of CT findings and outcome in COVID-19 patients with OP pattern.

## Materials and Methods

### Patients

This retrospective study was approved by the institutional review board and informed patients’ consent was waived with approval. 158 laboratory-confirmed patients with COVID-19 pneumonia who underwent chest CT scans between 22 January 2020 and 16 March 2020 were collected from eight hospitals in China. Among the patients, 75 were from Xi’an region; 18 were from Ankang region; 17 were from Hanzhong region; 10 patients were from Baoji region, and 38 patients were from Wuhan region. A case of COVID-19 was confirmed by a positive result on next-generation sequencing or real-time RT-PCR. The pulmonary lesions were considered to belong to OP pattern based on baseline CT: (1) peripheral predominantly GGO, consolidation or both, with subpleural or bronchovascular bundles distribution (Figure 4); (2) lobar involvement characterized by the total CT score less than or equal to 10 (Evaluation for total CT score detailed below). Unqualified CT images and CT characteristics unmatched with OP pattern were excluded.

According to clinical classification from preliminary diagnosis and treatment protocols for novel coronavirus pneumonia (7th edition) of the National Health Commission, China (13), all patients were assessed as mild, common, severe and critical types and categorized into non-severe (mild/common) and severe (severe/critical) groups. For mild patients, clinical symptoms are subtle and no pneumonia found on chest imaging. Patients with common type show symptoms such as fever and respiratory tract, and lung opacities on chest imaging. Patients with severe type should meet any of the following conditions: (1) respiratory distress, RR ≥30 beats / min; (2) resting blood oxygen saturation ≤93%; or (3) partial pressure of arterial blood oxygen (PaO2)/oxygen concentration (FiO2) ≤300 mmHg. Critical patients should meet one of the following criteria: (1) respiratory failure with mechanical ventilation; (2) shock; (3) other organ failure needing intensive care unit (ICU) treatment.

All the patients were diagnosed, isolated and hospitalized, which included initiation of antivirals, interferon, Chinese herbal medications, supplemental oxygen (13). The discharge criteria were: (1) body temperature returned to normal for greater than 3 days; (2) respiratory symptoms significantly improved; (3) pulmonary imaging showed obvious improvement in acute exudative lesions; (4) two consecutive negative COVID-19 nucleic acid tests at least 24 h apart. The discharged patients were recommended to quarantine for two weeks and then revisit the hospital with nucleic acid test and chest CT scan at 2 and 4 weeks after discharge (13). The pulmonary sequelae, i.e. complete absorption or residuals with linear opacities, and/or a few GGO with/without consolidation on revisit CT were evaluated. The disease course was defined as the interval from symptom onset to discharge.

### CT image acquisition

Chest CT scans were performed in 16- to 64-multidector CT scanners (Philips Brilliant 16, Philips Healthcare; GE LightSpeed 16, GE Healthcare; GE VCT LightSpeed 64, GE Healthcare; Somatom Sensation 64, Siemens Healthcare; Somatom AS, Siemens Healthcare; Somatom Spirit, Siemens Healthcare; GE Optima 680, GE Healthcare). The CT parameters included: 120 kVp of tube voltage, current intelligent control (auto mA) of 30-300 mA, and slice thickness/ slice interval of 0.6–1.5 mm.

### Image data collection and evaluation

Two experienced radiologists with respective 5 and 10 years of thoracic imaging experiences reviewed and described CT findings according to a peer-reviewed literature of viral pneumonia (9,11). The following terms were used: pure GGO; pure consolidation; GGO and consolidation; pure linear opacity; GGO and linear opacity; consolidation and linear opacity; GGO, consolidation and linear opacity; crazy paving pattern; reversed halo pattern. The pulmonary abnormalities involvement was quantitatively estimated by a semi-quantitative scoring system (14). Each of the five lung lobes was visually scored from 0 to 4 as: 0, no involvement; 1, <25% involvement; 2, 25%-49% involvement; 3, 50%-75% involvement; 4, >75% involvement. The sum of the individual lobar scores were the total CT scores, which ranged from 0 (no involvement) to 20 (maximum involvement).

CT findings were designated to three time groups based on the time from symptom onset to CT scan (day 0-7, day 8-14, day >14).

### Statistical Analysis

Continuous variables were presented as mean±standard deviation and the categorical variables were presented as the number and percentage of the total. Differences of CT findings across time groups (day 0-7, day 8-14, day >14), between non-severe and severe cases, between complete absorption and residuals groups were analyzed by Chi-square test, Fisher’s exact test, dependent sample t-test or Mann-Whitney U test as appropriate. Multiple comparisons were corrected by Bonferroni correction. Continuous variable on CT images, i.e. total CT score with significant difference in two-group comparison was further treated as categorical variable using an optimal threshold to maximize the Youden index of the receiver operating characteristic (ROC) analysis in discrimination of complete absorption vs. residuals groups.

All the statistical analyses were performed in the IBM SPSS Statistics Software (version 22; IBM, New York, USA). *P*<0.05 was considered statistically significant.

## Results

### Patient characteristics

Of 158 patients, 106(67.1%) were OP pattern, 3(1.9%) were with negative CT, and 49(31%) were unmatched with OP pattern (Figure 1). Of 106 OP pattern cases, 79(74.5%) were non-severe and 27(25.5%) were severe; 61(61.6%) were with elevated C-reactive protein and 42(40.0%) were with decreased lymphatic percentage (Table 1). The mean age was 48.0±15.4 years and showed no significant gender difference (male, 45[43.7%]; female, 61[56.3%]).

**Figure 1.**
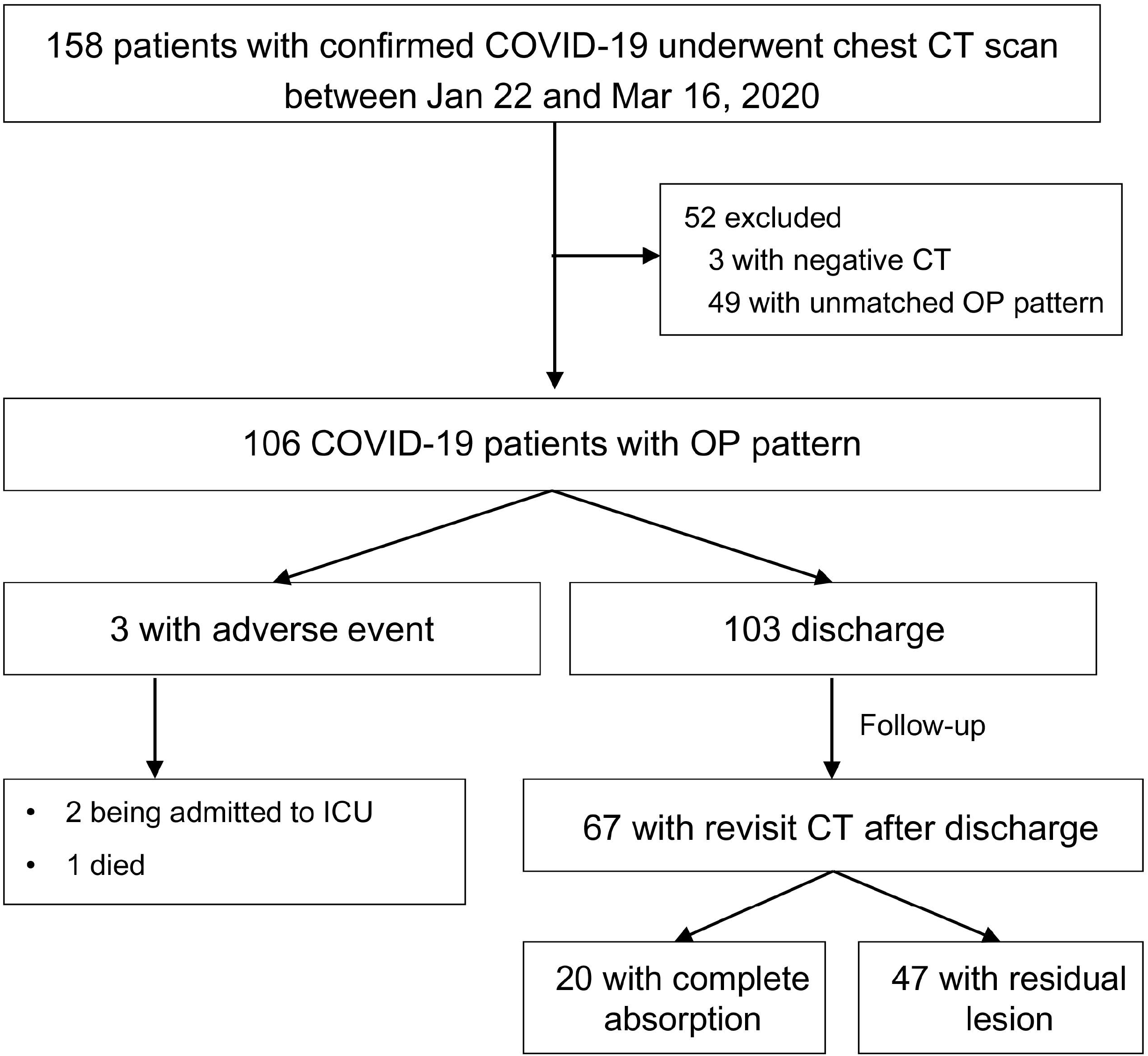
Patients recruitment flowchart. COVID-19 = coronavirus disease 2019, ICU = intensive care unit, OP = organizing pneumonia.

**Table 1.** Demographics, laboratory test and clinical outcome in COVID-19 patients with organizing pneumonia pattern.

| Characteristics | Patients<br>(n=106) |
| --- | --- |
| **Age (year)<sup>a</sup>** | 48.0±15.4 |
| **Male sex** | 45(43.7%) |
| **Disease severity** |  |
| Non-severe | 79(74.5%) |
| Severe | 27(25.5%) |
| **Comorbidity** | 38(35.8%) |
| **Exposure history** |  |
| Recent travel to Wuhan | 63(61.2%) |
| Contact with infected patient | 28(27.2%) |
| Unknown exposure | 12(11.7%) |
| **Initial symptom** |  |
| Fever | 91(88.4%) |
| Cough | 61(59.2%) |
| Expectoration | 21(20.4%) |
| Fatigue | 13(12.6%) |
| Chest tightness and/or breath shortness | 21(20.4%) |
| Pharyngalgia | 10(9.7%) |
| Muscle soreness | 7(6.8%) |
| Headache | 7(6.8%) |
| Nausea and/or vomiting | 1(1.0%) |
| Diarrhea | 4(3.9%) |
| No obvious symptoms | 2(1.9%) |
| **Laboratory test at admission<sup>b</sup>** |  |
| C-reactive protein (-,↑,↓) | 38(38.4%), 61(61.6%), 0(0) |
| Percentage of lymphocytes (-,↑,↓) | 60(57.1%), 3(2.9%), 42(40.0%) |
| Lymphocyte count (-,↑,↓) | 64(61.5%), 0(0), 40(38.5%) |
| Percentage of monocytes (-,↑,↓) | 75(72.1%), 26(25.0%), 3(2.9%) |
| White blood cell count (-,↑,↓) | 73(69.5%), 2(1.9%), 2(28.6%) |
| Alanine Aminotransferase (-,↑,↓) | 82(78.8%), 20(19.2%), 2(1.9%) |
| Aspartate Aminotransferase (-,↑,↓) | 82(78.8%), 21(20.2%), 1(1.0%) |
| Creatine kinase (-,↑,↓) | 84(83.2%), 7(6.9%), 10(9.9%) |
| Neutrophilic granulocyte percentage (-,↑,↓) | 69(65.7%), 26(24.8%), 10(9.5%) |
| Hemoglobin (-,↑,↓) | 79(76.0%), 6(5.8%), 19(18.3) |
| **Clinical outcome** |  |
| Discharge | 103(97.2%) |
| Admission to ICU | 2(1.9%) |
| Death | 1(0.9%) |

Note: Unless otherwise indicated, data are reported as the number of patients, with percentages in parentheses.
<sup>a</sup>, data were reported as the mean±standard derivation.
<sup>b</sup>, -, ↑, ↓ represent within, above, and below normal ranges of laboratory results, respectively. Normal ranges of C-reactive protein, percentage of lymphocytes, lymphocyte count, percentage of monocytes, white blood cell count, ALT, AST, creatine kinase, neutrophilic granulocyte percentage and hemoglobin were 0-10 mg/L, 20-50%, $1.10-3.20 \times 10^9/L$ , 3.0-10.0%, $3.5-9.5 \times 10^9/L$ , 7-40 U/L, 13-35 U/L, 40-200 U/L, 40-75%, 115-150 g/L, respectively.

Of 106 OP pattern cases, 103(97.2%) were discharged, 2(1.9%) were admitted to ICU, 1(0.9%) died. Median times from symptom onset to discharge, to ICU admission, to death were 25 (range, 8-50) days, 24 days, 28 days, respectively. Of 67 patients with revisit CT after discharge, 20(29.9%) had complete resolution with a median interval of 38 (range, 30-53) days between symptom onset and revisit CT after discharge. A total of 340 CT scans were obtained from 106 patients. The average number of CT scans was 3 (range, 1-8).

### CT findings of OP pattern in COVID-19

Of 1285 lesions in 106 patients, pure GGO (32.2%) was the predominant CT finding, followed by the mixed GGO and consolidation (21.2%), mixed GGO, consolidation and linear opacity (17.7%). Pure linear opacity, reversed halo signs and crazy paving were rare, accounting for 1.9%, 2.2% and 2.0% respectively. Mean total CT score was 5.1±2.8. For number of involved lobes, the lesions were mostly located in bilateral lower lobes (right, 24.7%; left, 22.8%) (Table S1).

Significant differences of pure GGO, pure consolidation, pure linear opacity, mixed consolidation and linear opacity, involvement of lung lobes and total CT score were found among the three time groups (day 0-7, day 8-14, day >14) (all *P*<0.05). From day 0-7 to day 8-14, the percentage of pure GGO significantly decreased (41.4% vs. 30.6%, *P*=0.002); despite no significance with Bonferroni correction, involvement of lung lobes and CT score remarkably increased. From day 8-14 to day >14, the percentage of pure liner opacity significantly increased (1.0% vs. 3.3%, *P*=0.03) (Figure 2, Table S2).

**Figure 2.**
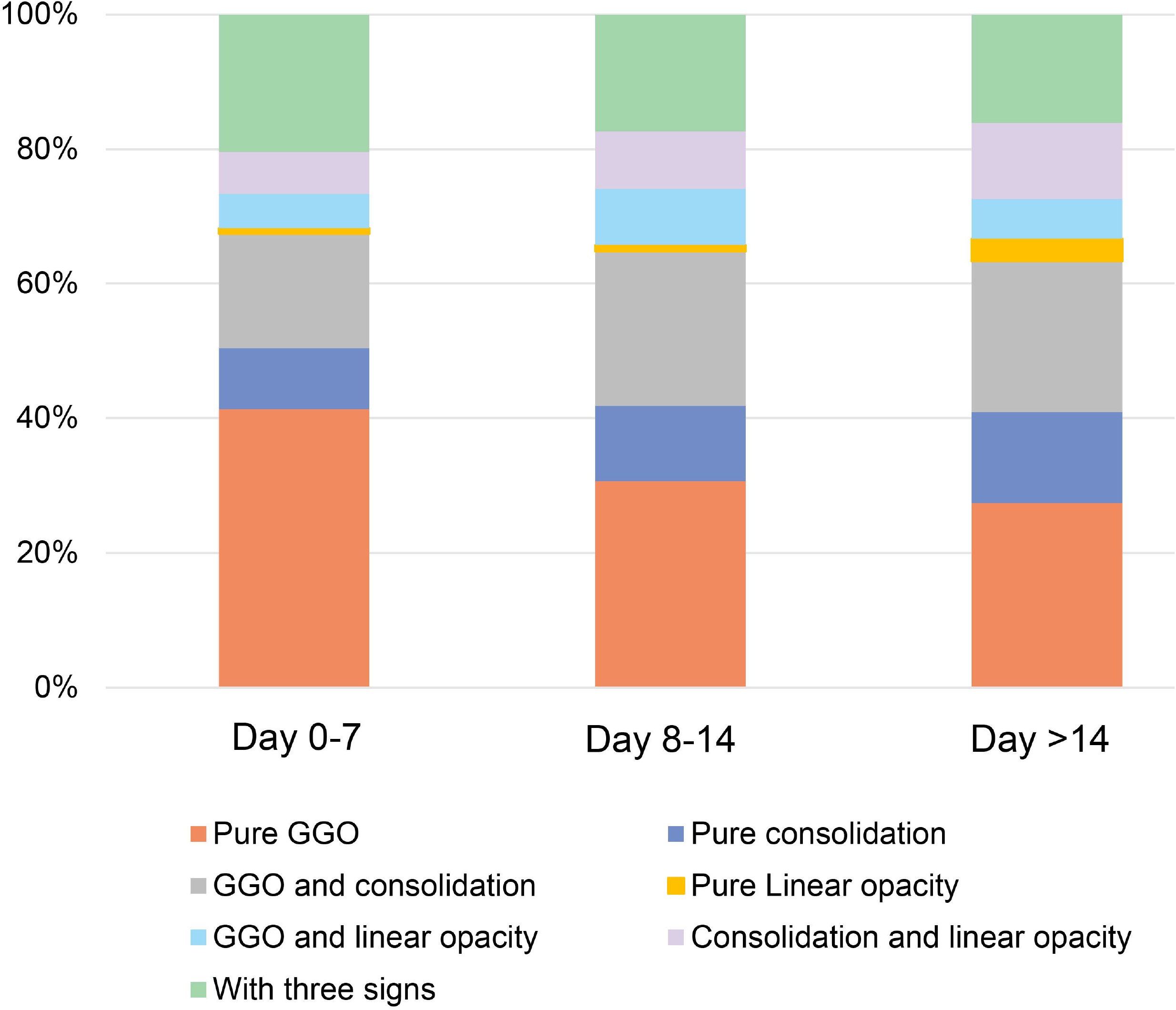
The evolution of CT findings across the three time groups (day 0-7, day 8-14, day >14) in COVID-19 patients with organizing pneumonia pattern. GGO = ground glass opacity; with three signs = GGO, consolidation and linear opacity.

### Comparisons of clinical and CT findings between non-severe and severe cases

Significant differences between non-severe and severe cases were found in terms of age (44.8±13.5 vs. 59.9±12.5 years, *P*<0.001), presence of comorbidity (27.5% vs. 59.3%, *P*=0.006), symptom of chest tightness and/or breath shortness (13.9% vs. 40.7%, *P*=0.003) and decreased lymphocyte count (30.8% vs. 61.5%, *P*=0.005) (Table 2).

**Table 2.**
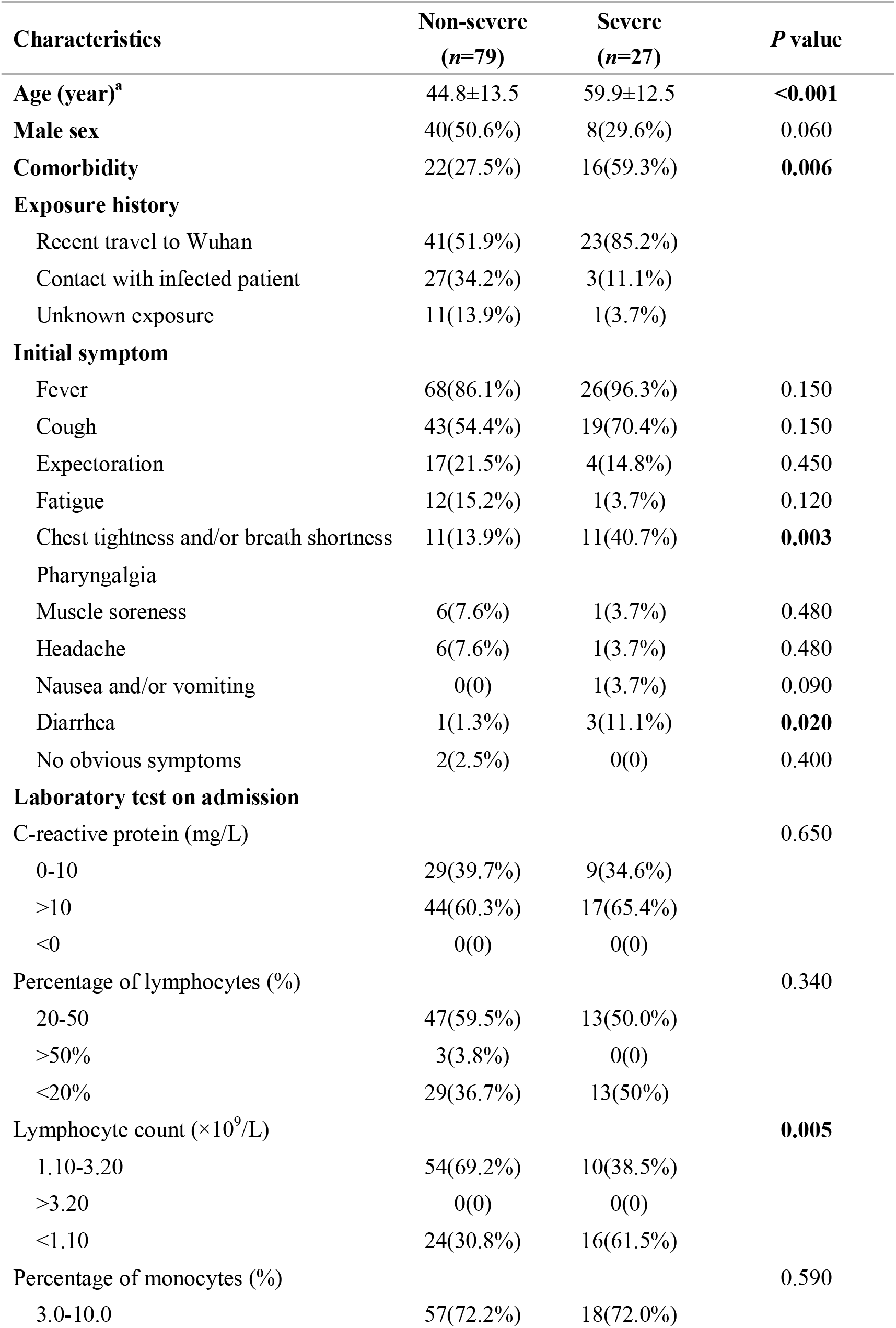
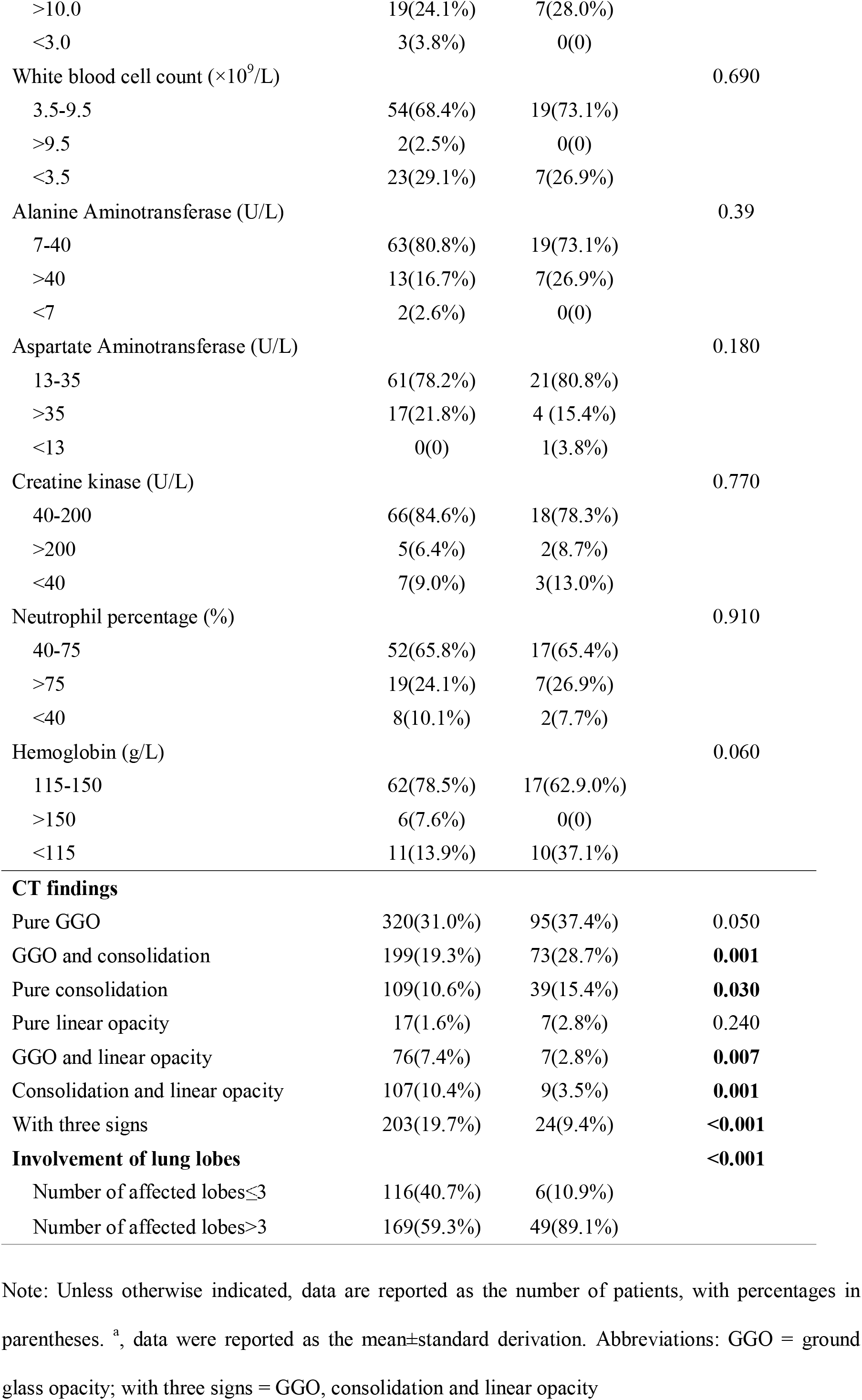
Comparisons of clinical characteristics and CT findings between non-severe and severe groups in COVID-19 patients with organizing pneumonia.

| Characteristics | Non-severe<br>(n=79) | Severe<br>(n=27) | P value |
| --- | --- | --- | --- |
| Age (year)<sup>a</sup> | 44.8±13.5 | 59.9±12.5 | <0.001 |
| Male sex | 40(50.6%) | 8(29.6%) | 0.060 |
| Comorbidity | 22(27.5%) | 16(59.3%) | **0.006** |
| Exposure history |  |  |  |
| Recent travel to Wuhan | 41(51.9%) | 23(85.2%) |  |
| Contact with infected patient | 27(34.2%) | 3(11.1%) |  |
| Unknown exposure | 11(13.9%) | 1(3.7%) |  |
| Initial symptom |  |  |  |
| Fever | 68(86.1%) | 26(96.3%) | 0.150 |
| Cough | 43(54.4%) | 19(70.4%) | 0.150 |
| Expectoration | 17(21.5%) | 4(14.8%) | 0.450 |
| Fatigue | 12(15.2%) | 1(3.7%) | 0.120 |
| Chest tightness and/or breath shortness | 11(13.9%) | 11(40.7%) | **0.003** |
| Pharyngalgia |  |  |  |
| Muscle soreness | 6(7.6%) | 1(3.7%) | 0.480 |
| Headache | 6(7.6%) | 1(3.7%) | 0.480 |
| Nausea and/or vomiting | 0(0) | 1(3.7%) | 0.090 |
| Diarrhea | 1(1.3%) | 3(11.1%) | **0.020** |
| No obvious symptoms | 2(2.5%) | 0(0) | 0.400 |
| Laboratory test on admission |  |  |  |
| C-reactive protein (mg/L) |  |  | 0.650 |
| 0-10 | 29(39.7%) | 9(34.6%) |  |
| >10 | 44(60.3%) | 17(65.4%) |  |
| <0 | 0(0) | 0(0) |  |
| Percentage of lymphocytes (%) |  |  | 0.340 |
| 20-50 | 47(59.5%) | 13(50.0%) |  |
| >50% | 3(3.8%) | 0(0) |  |
| <20% | 29(36.7%) | 13(50%) |  |
| Lymphocyte count (×10<sup>9</sup>/L) |  |  | **0.005** |
| 1.10-3.20 | 54(69.2%) | 10(38.5%) |  |
| >3.20 | 0(0) | 0(0) |  |
| <1.10 | 24(30.8%) | 16(61.5%) |  |
| Percentage of monocytes (%) |  |  | 0.590 |
| 3.0-10.0 | 57(72.2%) | 18(72.0%) |  |
| >10.0 | 19(24.1%) | 7(28.0%) |  |
| <3.0 | 3(3.8%) | 0(0) |  |
| White blood cell count ($\times 10^9/L$) | | | 0.690 |
| 3.5-9.5 | 54(68.4%) | 19(73.1%) |  |
| >9.5 | 2(2.5%) | 0(0) |  |
| <3.5 | 23(29.1%) | 7(26.9%) |  |
| Alanine Aminotransferase (U/L) |  |  | 0.39 |
| 7-40 | 63(80.8%) | 19(73.1%) |  |
| >40 | 13(16.7%) | 7(26.9%) |  |
| <7 | 2(2.6%) | 0(0) |  |
| Aspartate Aminotransferase (U/L) |  |  | 0.180 |
| 13-35 | 61(78.2%) | 21(80.8%) |  |
| >35 | 17(21.8%) | 4 (15.4%) |  |
| <13 | 0(0) | 1(3.8%) |  |
| Creatine kinase (U/L) |  |  | 0.770 |
| 40-200 | 66(84.6%) | 18(78.3%) |  |
| >200 | 5(6.4%) | 2(8.7%) |  |
| <40 | 7(9.0%) | 3(13.0%) |  |
| Neutrophil percentage (%) |  |  | 0.910 |
| 40-75 | 52(65.8%) | 17(65.4%) |  |
| >75 | 19(24.1%) | 7(26.9%) |  |
| <40 | 8(10.1%) | 2(7.7%) |  |
| Hemoglobin (g/L) |  |  | 0.060 |
| 115-150 | 62(78.5%) | 17(62.9.0%) |  |
| >150 | 6(7.6%) | 0(0) |  |
| <115 | 11(13.9%) | 10(37.1%) |  |
| **CT findings** |  |  |  |
| Pure GGO | 320(31.0%) | 95(37.4%) | 0.050 |
| GGO and consolidation | 199(19.3%) | 73(28.7%) | **0.001** |
| Pure consolidation | 109(10.6%) | 39(15.4%) | **0.030** |
| Pure linear opacity | 17(1.6%) | 7(2.8%) | 0.240 |
| GGO and linear opacity | 76(7.4%) | 7(2.8%) | **0.007** |
| Consolidation and linear opacity | 107(10.4%) | 9(3.5%) | **0.001** |
| With three signs | 203(19.7%) | 24(9.4%) | **<0.001** |
| **Involvement of lung lobes** |  |  | **<0.001** |
| Number of affected lobes$\leq 3$ | 116(40.7%) | 6(10.9%) | |
| Number of affected lobes$>3$ | 169(59.3%) | 49(89.1%) | |

Note: Unless otherwise indicated, data are reported as the number of patients, with percentages in parentheses. <sup>a</sup>, data were reported as the mean±standard derivation. Abbreviations: GGO = ground glass opacity; with three signs = GGO, consolidation and linear opacity

There were 1031 and 254 lesions in non-severe and severe cases, respectively. The percentage of mixed GGO and consolidation and pure consolidation were significantly lower in non-severe group than those in severe group (19.3% vs. 28.7%, *P*=0.001; 10.6% vs. 15.4%, *P*=0.030), while percentage of mixed GGO and linear opacity, mixed consolidation and linear opacity, mixed three signs were significantly higher in non-severe than severe groups (7.4% vs. 2.8%, *P*=0.007; 10.4% vs. 3.5%, *P*=0.001; 19.7% vs. 29.4%, *P*<0.001). The number of affected lobes>3 and total CT score were significantly lower in non-severe than severe groups (59.3% vs. 89.1%, *P*<0.001; 4.7±2.5 vs. 7.5±2.9, *P*<0.001) (Table 2).

### Comparisons of clinical and CT findings between complete absorption and residual groups

Significant difference between complete absorption and residuals groups were found in age (34.9±9.0 vs. 47.9±13.7, *P*=0.001), elevated C-reactive protein (31.6% vs. 70.5%, *P*=0.009) and neutrophil percentage (10.0% vs. 37.0%, *P*=0.020) (Table 3). At day 0-7, 81 and 185 lesions were found on CT in complete absorption and residual groups, respectively. Percentage of pure consolidation was significantly higher in complete absorption than residuals groups (14.8% vs. 5.9%, *P*=0.02) (Figure 3, Table S3).

**Figure 3.**
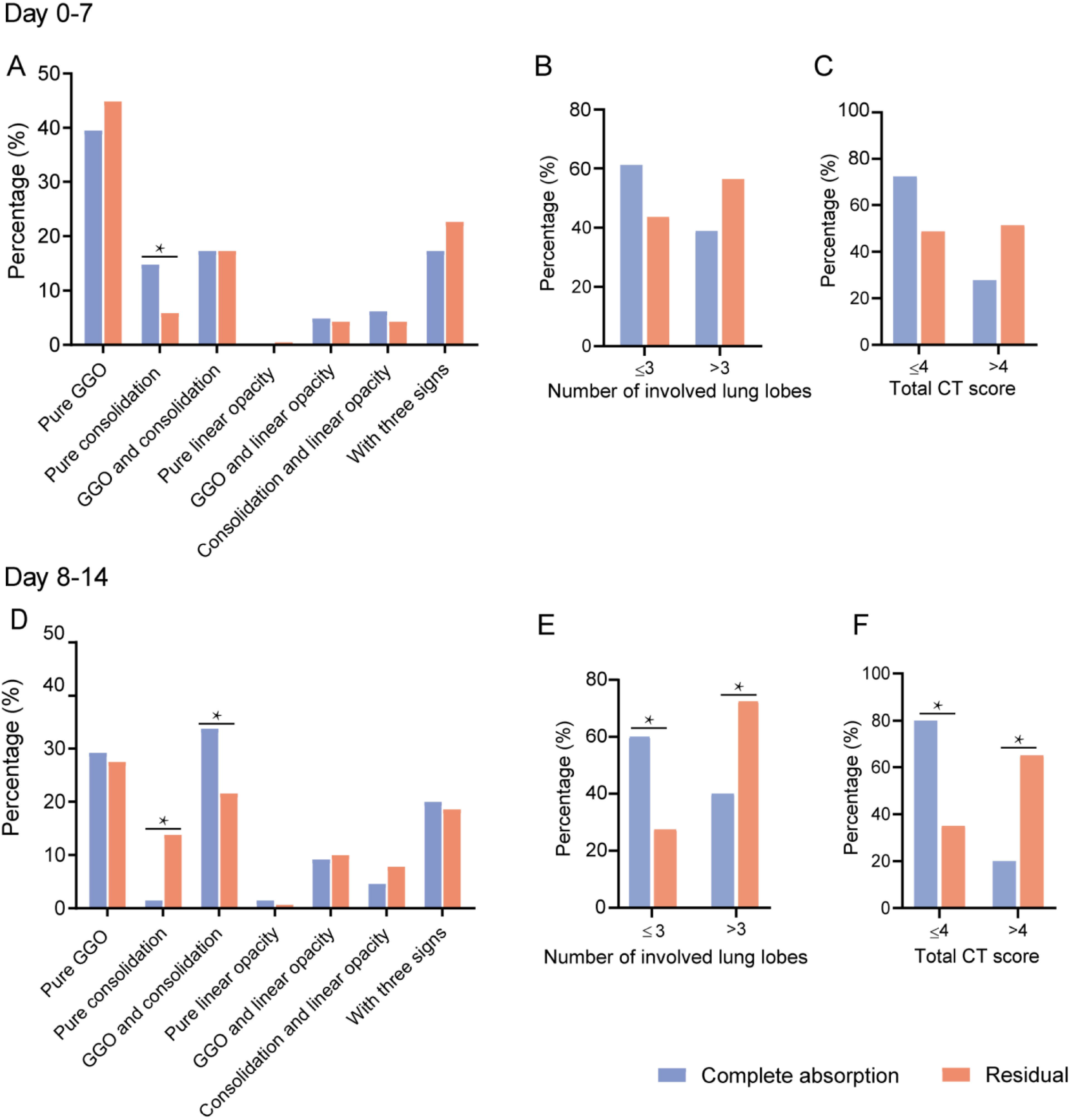
Comparisons of CT findings between patients with complete absorption and residuals after discharge. GGO = ground glass opacity; with three signs = GGO, consolidation and linear opacity. *, *P<* 0.05 indicated significant differences.

**Figure 4.**
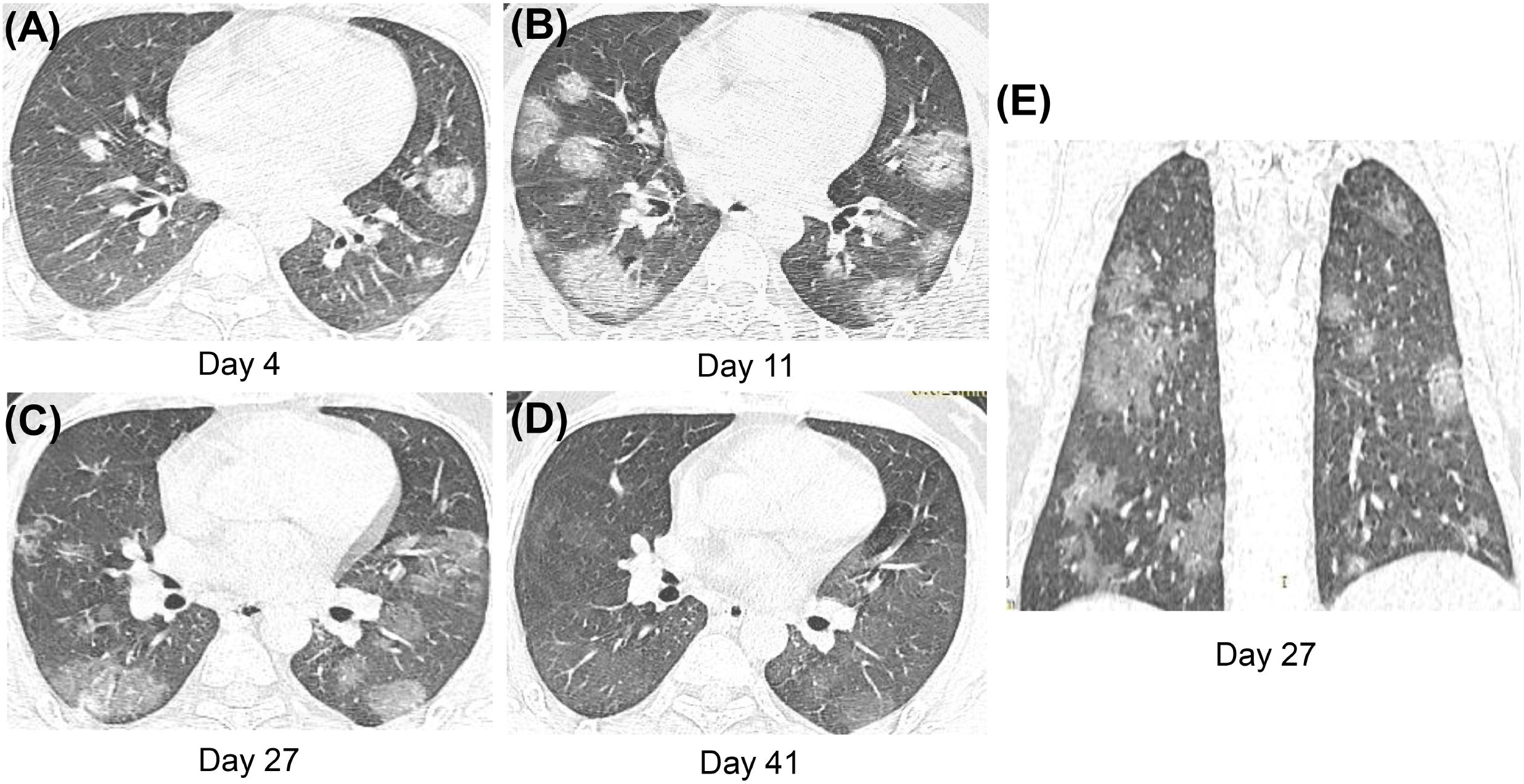
A 66-year-old man who had been to Wuhan had fever for 4 days and discharged at day 28 after symptom onset. (A) CT obtained on admission at day 4 after symptom onset shows multiple subpleural consolidation. (B) CT on day 11 shows progression with increased number and size of lung lesions. (C, E) CT on day 27 shows absorption with decreased density of the lesions. (D) CT on day 41 shows almost complete absorption of pulmonary lesions.

A total of 65 and 269 lesions in complete absorption and residuals groups were found at day 8-14. Significant differences between complete absorption and residuals groups were found in percentage of mixed GGO and consolidation (33.8% vs. 21.6%, *P*=0.04), percentage of pure consolidation (1.5% vs. 13.8%, *P*=0.01), percentage of affected lobe number>3 (40.0% vs. 72.5%, *P*=0.03) (Figure 3, Table S3). In addition, ROC curve analysis estimated a cutoff CT score of 4 to discriminate the complete absorption from residuals groups at day 8-14 (AUC=0.732, *P*=0.006) (Figure S1). Significantly higher percentage of CT score >4 was found in residuals than complete absorption groups (65.0% vs. 20.0%, *P*=0.001) (Figure 3, Table S3).

**Table 3.** Comparisons of demographics and laboratory test between complete absorption and residual groups in COVID-19 patients with organizing pneumonia.

| Characteristics | Complete absorption<br>(n=20) | Residual<br>(n=47) | P value |
| --- | --- | --- | --- |
| **Age (year)<sup>a</sup>** | 34.9±9.0 | 47.9±13.7 | **0.001** |
| **Male sex** | 11(55.0%) | 9(45.0%) | 0.710 |
| **Comorbidity** | 3(15.0%) | 18(38.3%) | 0.110 |
| **Exposure history** |  |  | 0.410 |
| Recent travel to Wuhan | 13(65.0%) | 22(47.8%) |  |
| Contact with infected patient | 5(25.0%) | 15(32.6%) |  |
| Unknown exposure | 2(10.0%) | 9(19.6%) |  |
| **Initial symptom** |  |  |  |
| Fever | 17(85.0%) | 42(91.3%) | 0.450 |
| Cough | 8(40.0%) | 27(58.7%) | 0.160 |
| Expectoration | 3(15.0%) | 15(32.6%) | 0.240 |
| Fatigue | 3(15.0%) | 7(15.2%) | 0.980 |
| Chest tightness and/or breath | 2(10.0%) | 6(13.0%) | 0.990 |
| Pharyngalgia | 2(10.0%) | 7(15.2%) | 0.570 |
| Muscle soreness | 2(10.0%) | 5(10.9%) | 0.990 |
| Headache | 1(5.0%) | 4(8.7%) | 0.990 |
| Nausea and/or vomiting | 0(0) | 1(2.2%) | 0.990 |
| Diarrhea | 0(0) | 2(4.3%) | 0.870 |
| No obvious symptoms | 1(5.0%) | 1(2.2%) | 0.990 |
| **Laboratory test on admission** |  |  |  |
| **C-reactive protein (mg/L)** |  |  | **0.009** |
| 0-10 | 13(68.4%) | 13(29.5%) |  |
| >10 | 6(31.6%) | 31(70.5%) |  |
| <0 | 0(0) | 0(0) |  |
| **Percentage of lymphocytes (%)** |  |  | 0.560 |
| 20-50 | 12(60.0%) | 23(50.0%) |  |
| >50 | 1(5.0%) | 1(2.2%) |  |
| <20 | 7(35.0%) | 22(47.8%) |  |
| **Lymphocyte count (×10<sup>9</sup>/L)** |  |  | 0.140 |
| 1.10-3.20 | 15(75.0%) | 25(55.6%) |  |
| >3.20 | 0(0) | 0(0) |  |
| <1.10 | 5(25.0%) | 20(44.4%) |  |
| Percentage of monocytes (%) |  |  | 0.630 |
| 3.0-10.0 | 13(65.0%) | 35(76.1%) |  |
| >10.0 | 6(30.0%) | 9(19.6%) |  |
| <3.0 | 1(5.0%) | 2(4.3%) |  |
| White blood cell count ($\times 10^9/L$) | | | 0.710 |
| 3.5-9.5 | 14(70.0%) | 30(65.2%) |  |
| >9.5 | 1(5.0%) | 1(2.2%) |  |
| <3.5 | 5(25.0%) | 15(32.6%) |  |
| Alanine Aminotransferase (U/L) |  |  | 0.540 |
| 7-40 | 17(85.0%) | 34(75.6%) |  |
| >40 | 3(15.0%) | 9(20.0%) |  |
| <7 | 0(0) | 2(4.4%) |  |
| Aspartate Aminotransferase (U/L) |  |  | 0.840 |
| 13-35 | 16(80.0%) | 35(77.8%) |  |
| >35 | 4(20.0%) | 10(22.2%) |  |
| <13 | 0(0) | 0(0) |  |
| Creatine kinase (U/L) |  |  | 0.130 |
| 40-200 | 15(75.0%) | 39(86.7%) |  |
| >200 | 1(5.0%) | 4(8.9%) |  |
| <40 | 4(20.0%) | 2(4.4%) |  |
| Neutrophil percentage (%) |  |  | **0.020** |
| 40-75 | 14(70.0%) | 27(58.7%) |  |
| >75 | 2(10.0%) | 17(37.0%) |  |
| <40 | 4(20.0%) | 2(4.3%) |  |
| Hemoglobin (g/L) |  |  | 0.050 |
| 115-150 | 18(90.0%) | 36(78.3%) |  |
| >150 | 2(10.0%) | 1(2.2%) |  |
| <115 | 0(0) | 9(19.6%) |  |

Note: Unless otherwise indicated, data are reported as the number of patients, with percentages in parentheses.
<sup>a</sup>, data were reported as the mean ± standard derivation.

## Discussion

With regard to the most common OP pattern in COVID-19, this study systematically described the clinical characteristics and time-dependent evolution of CT findings, as well as later outcome of patients. Results indicated that 74.5% of OP cases were non-severe and 97.1% cases had good prognosis with recovery. As for pulmonary resolution, approximately one-third of OP cases had complete absorption of lesions during day 30-53 after symptom onset while those with increased percentages of consolidation, number of involved lung lobe >3, and CT score >4 at week 2 after onset are prone to have pulmonary residuals.

Being consistent with previous finding of COVID-19 (14), the dominant finding in OP pattern was the presence of GGO, followed by mixed GGO and consolidation, with peripheral and lower lobes distribution. In addition, time-dependent evolution indicated that percentage of GGO decreased while that of mixed GGO and consolidation increased from 1 to 2 weeks after onset, and linear opacity increased from 2 to 3 weeks after onset. These findings were in accordance with prior reports regarding the radiological aggravation (≤2 weeks) and improvement (>2 weeks) in COVID-19 (9,10). A systematic review in COVID-19 also observed that GGO turned into extensive consolidation with greatest severity at around day 10 after onset and consolidation gradually resolved after 2 weeks (15). Histopathologically, alveolar epithelium injured, inflammatory component, fibrin deposition and matrix leaked and coagulated in early and advanced stage and then gradually receded and resorbed during the late disease course (16), which may explain the primary CT findings in OP pattern.

In COVID-19, 74.5% of OP pattern cases were non-severe which was in agreement with the mild degree of lung injury in most OP cases (17). In addition, severe cases had older age, more prevalence of comorbidity, and decreased lymphocyte count than non-severe cases. Besides, more prevalence of mixed GGO and consolidation, and pure consolidation with higher lung severity were found in severe cases, whereas non-severe cases showed more prevalence of linear opacity. This may imply a progressive pulmonary involvement in severe cases while a reparative process in non-severe cases. It may be such facts that led to the longer course of disease from symptom onset to discharge in severe than non-severe cases (36.5±9.9 vs. 23.9±7.4 days, *P*<0.001).

As for clinical outcome, most of OP pattern cases showed good prognosis with discharge. This resembled the previous study of OP (17). It is noting that for 3 patients with adverse outcomes, they had progressively diffuse GGO and consolidation with interlobular septal thickening in both lungs within the first week after onset. This may be linked to a fast progression from OP to diffuse alveolar damage (18). Among the discharged patients, those with increased percentages of consolidation, number of involved lung lobe >3, and CT score >4 at week 2 after onset were prone to have pulmonary residuals. What we observed during the first two weeks was probably correlated with the underlying organizing process of lung injury (7). Prior study found that extensive consolidation as well as increased CT score may suggest the disease progression (11,12). In this regard, cases with extensive consolidation and progressive lung involvement may have a protracted disease course of lesion absorption. Similarly, radiological sequelae were also observed in Severe Acute Respiratory Syndrome (SARS) and Middle East Respiratory Syndrome (MERS). In details, radiological sequelae with impaired pulmonary function was found in MERS at the 1-year after infection (19). Antonio et al. found that some SARS patients showed residual abnormalities on CT with average interval of 18 days after discharge, which was similar to our study (20). Note that radiological sequelae from SARS and MERS may suggest the repaired lung function (20). Differently, slighter residuals mainly presenting with linear opacities was found in OP pattern of COVID-19. Beyond, elevated C-reactive protein and neutrophil percentage may indicate the state of tissue injury and/or inflammation (21). Previous study indicated that continuous high levels of C-reactive protein in respiratory infections increases the risk of progression to a critical disease state (22). In this regard, elevated C-reactive protein and neutrophil percentage may be predictive of radiological sequelae. Although OP cases of COVID-19 had a favourable prognosis, early monitoring and detection of adverse outcomes and radiological sequelae would contribute to the early intervention for those with potential risks to fibrosis, respiratory failure and death (23).

There were several limitations in this study. First was the relatively small sample and retrospective nature. A larger sample is required to further validate the findings regarding OP of COVID-19. Second, the follow-up period for patients is relatively short and many of residual lesions on CT may be reversible, a long-term follow-up in conjunction with lung function tests would help to further clarify the evolution of residual lesions and its relations with lung function. Third, although pathological evidence is scarce, highly resembled CT features of OP were used to define the OP pattern of COVID-19 in this study. Pathological studies are still needed to validate the OP of COVID-19.

In conclusion, as a most common pattern of COVID-19, majority of OP cases were mild or common and had good prognosis. Approximately one-third of OP cases had complete absorption of lesions during 1-2 month after symptom onset while those with increased frequency of pure consolidation, number of involved lung lobe >3, and CT score >4 at week 2 after symptom onset were prone to have pulmonary residuals.

## Data Availability

Data is not available

## References

1. Zhu N, Zhang D, Wang W, Li X, Yang B, Song J, et al. A Novel Coronavirus from Patients with Pneumonia in China, 2019. N. Engl. J. Med. 2020;382(8):727–733.

2. Huang C, Wang Y, Li X, Ren L, Zhao J, Hu Y, et al. Clinical features of patients infected with 2019 novel coronavirus in Wuhan, China. Lancet. 2020 Feb 15; 395(10223):497–506.

3. Guan W, Ni Z, Hu Y, Liang W, Ou C, He J, et al. Clinical characteristics of 2019 novel coronavirus infection in China. N Engl J Med. 2020 Apr 30;382 (18): 1708-1720.

4. WHO. Coronavirus disease 2019. 2020. https://www.who.int/emergencies/diseases/novel-coronavirus-2019/situation-reports. Accessed April 29, 2020.

5. Kanne JP, Little BP, Chung JH, Elicker BM, Ketai LH. Essentials for Radiologists on COVID-19: An Update—Radiology Scientific Expert Panel. Radiology. 2020 Feb 27: 200527.

6. Simpson S, Kay FU, Abbara S, Bhalla S, Chung JH, Chung M, et al. Radiological Society of North America Expert Consensus Statement on Reporting Chest CT Findings Related to COVID-19. Endorsed by the Society of Thoracic Radiology, the American College of Radiology, and RSNA. J Thorac Imaging. 2020 Apr 28 [Epub]. doi: 10.1097/RTI.0000000000000524.

7. Kligerman SJ, Franks TJ, Galvin JR. From the Radiologic Pathology Archives: Organization and fibrosis as a response to lung injury in diffuse alveolar damage, organizing pneumonia, and acute fibrinous and organizing pneumonia. Radiographics. 2013;33(7):1951–1975.

8. Chen J, Qi T, Liu L, Ling Y, Qian Z, Li T, et al. Clinical progression of patients with COVID-19 in Shanghai, China. J Infect. 2020 May;80(5): e1-e6.

9. Shi H, Han X, Jiang N, Cao Y, Alwalid O, Gu J, et al. Radiological findings from 81 patients with COVID-19 pneumonia in Wuhan, China: a descriptive study. Lancet Infect Dis. 2020 Apr;20(4):425–434.

10. Pan F, Ye T, Sun P, Gui S, Liang B, Li L, et al. Time Course of Lung Changes On Chest CT During Recovery From 2019 Novel Coronavirus (COVID-19) Pneumonia. Radiology. 2020 Feb 13:200370 [Epub]. doi: 10.1148/radiol.2020200370.

11. Wang Y, Dong C, Hu Y, Li C, Ren Q, Zhang X, et al. Temporal Changes of CT Findings in 90 Patients with COVID-19 Pneumonia: A Longitudinal Study. Radiology. 2020 Mar 19:200843 [Epub]. doi: 10.1148/radiol.2020200843.

12. Zu ZY, Jiang M Di, Xu PP, Chen W, Ni QQ, Lu GM, et al. Coronavirus Disease 2019 (COVID-19): A Perspective from China. Radiology. 2020 Feb 21:200490 [Epub]. doi: 10.1148/radiol.2020200490.

13. National Health Commission of the People’s Republic of China. Diagnosis and Treatment of Pneumonia Caused by 2019-nCoV (version 7). http://www.gov.cn/zhengce/zhengceku/2020-03/04/content_5486705.htm. Accessed April 29, 2020.

14. Chung M, Bernheim A, Mei X, Zhang N, Huang M, Zeng X, et al. CT imaging features of 2019 novel coronavirus (2019-NCoV). Radiology. 2020 Apr;295(1):202–207.

15. Salehi S, Abedi A, Balakrishnan S, Gholamrezanezhad A. Coronavirus Disease 2019 (COVID-19): A Systematic Review of Imaging Findings in 919 Patients. AJR Am J Roentgenol. 2020 Mar 14: 1-7.

16. Roberton BJ, Hansell DM. Organizing pneumonia: A kaleidoscope of concepts and morphologies. Eur Radiol. 2011 Nov;21(11):2244–54.

17. Drakopanagiotakis F, Paschalaki K, Abu-Hijleh M, Aswad B, Karagianidis N, Kastanakis E, et al. Cryptogenic and secondary organizing pneumonia: Clinical presentation, radiographic findings, treatment response, and prognosis. Chest. 2011 Apr; 139(4):893–900.

18. Xu Z, Shi L, Wang Y, Zhang J, Huang L, Zhang C, et al. Pathological findings of COVID-19 associated with acute respiratory distress syndrome. Lancet Respir Med. 2020 Apr;8(4):420–422.

19. Park W, Jun K, Kim N-H, Choi J-P, Rhee J-Y, Cheon S, et al. Correlation between Pneumonia Severity and Pulmonary Complications in Middle East Respiratory Syndrome. J Korean Med Sci. 2018 May 10;33(24): e169.

20. Antonio GE, Wong KT, Hui DSC, Wu A, Lee N, Yuen EHY, et al. Thin-Section CT in Patients with Severe Acute Respiratory Syndrome Following Hospital Discharge: Preliminary Experience. Radiology. 2003 Sep;228(3):810–5.

21. Jenne CN, Liao S, Singh B. Neutrophils: multitasking first responders of immunity and tissue homeostasis. Cell Tissue Res. 2018 Mar;371(3):395–397.

22. Clyne B, Olshaker JS. The C-reactive protein. J Emerg Med. 1999 Nov-Dec;17(6):1019–25.

23. Cordier J-F. Cryptogenic organising pneumonia. Eur Respir J. 2006 Aug;28(2):422–46.

